# Emulation of epidemics via Bluetooth-based virtual safe virus spread: experimental setup, software, and data

**DOI:** 10.1101/2022.03.31.22273262

**Authors:** Azam Asanjarani, Aminath Shausan, Keng Chew, Thomas Graham, Shane G. Henderson, Hermanus M. Jansen, Kirsty R. Short, Peter G. Taylor, Aapeli Vuorinen, Yuvraj Yadav, Ilze Ziedins, Yoni Nazarathy

**Affiliations:** Faculty of Science, The University of Auckland, Auckland, State, New Zealand; School of Mathematics and Physics, The University of Queensland, Brisbane, Queensland, Australia; School of Chemistry and Molecular Biosciences, The University of Queensland, Brisbane, Queensland, Australia; School of Operations Research and Information Engineering, Cornell University, Ithaca, NY, USA; Department of Applied Mathematics, Delft University of Technology, Mekelweg 4, 2628CD Delft, The Netherlands; Mathematics and Statistics, The University of Melbourne, Melbourne, Victoria, Australia; Department of Industrial Engineering and Operations Research, Columbia University, New York, NY, USA; Mechanical Engineering Department, Indian Institute of Technology Delhi, New Delhi, Delhi, India

## Abstract

We describe an experimental setup and a currently running experiment for evaluating how physical interactions over time and between individuals affect the spread of epidemics. Our experiment involves the voluntary use of the Safe Blues Android app by participants at The University of Auckland (UoA) City Campus in New Zealand. The app spreads multiple virtual safe virus strands via Bluetooth depending on the social and physical proximity of the subjects. The evolution of the virtual epidemics is recorded as they spread through the population. The data is presented as a real-time (and historical) dashboard. A simulation model is applied to calibrate strand parameters. Participants’ locations are not recorded, but participants are rewarded based on the duration of participation within a geofenced area, and aggregate participation numbers serve as part of the data. Once the experiment is complete, the data will be made available as an open-source anonymized dataset.

This paper outlines the experimental setup, software, subject-recruitment practices, ethical considerations, and dataset description. The paper also highlights current experimental results in view of the lockdown that started in New Zealand at 23:59 on August 17, 2021. The experiment was initially planned in the New Zealand environment, expected to be free of COVID and lockdowns after 2020. However, a COVID Delta strain lockdown shuffled the cards and the experiment is currently extended into 2022.

**Author summary:** In this paper, we describe the Safe Blues Android app experimental setup and a currently running experiment at the University of Auckland City Campus. This experiment is designed to evaluate how physical interactions over time and between individuals affect the spread of epidemics.

The Safe Blues app spreads multiple virtual safe virus strands via Bluetooth based on the subjects’ unobserved social and physical proximity. The app does not record the participants’ locations, but participants are rewarded based on the duration of participation within a geofenced area, and aggregate participation numbers serve as part of the data. When the experiment is finished, the data will be released as an open-source anonymized dataset.

The experimental setup, software, subject recruitment practices, ethical considerations, and dataset description are all described in this paper. In addition, we present our current experimental results in view of the lockdown that started in New Zealand at 23:59 on August 17, 2021. The information we provide here may be useful to other teams planning similar experiments in the future.

## Introduction

The COVID-19 pandemic is the most significant global event of the 21st century to date. In response to the pandemic, multiple solutions have been and are still being developed and deployed, including vaccines and contact tracing technologies. As part of this effort, various initiatives that integrate digital health and “AI systems” (artificial intelligence for pandemics) are being thought out. A key initiative includes measuring the spread of pathogens as well as the level of physical human contact. The Safe Blues project is one such idea, where virtual safe virus-like tokens are spread between cellular phones in an attempt to mimic biological virus spread for purposes of measurement and analysis, while respecting the privacy and safety of the population.

Much COVID-19 data is being gathered by contact-tracing apps to aid in identifying infected people or their contacts. However, there can be a time lag of 1 to 2 weeks between being infected and being diagnosed as positive with the result that data obtained in this way is always lagging and biased. Also, asymptomatic cases who may have already spread the virus to others are frequently missed by such methods. Data delays and bias make it difficult for public health officials and others who want to use the data to implement timely mitigation measures. Our approach, on the other hand, is specifically designed to make inferences about the global characteristics of an epidemic in real-time, allowing governments to implement relevant mitigation measures in a timely fashion.

Safe Blues, introduced in [1, 2], works by spreading virtual ‘virus-like’ tokens, which we call *strands*. The strands can be of Susceptible-Exposed-Infectious-Removed (SEIR), Susceptible-Infectious-Removed (SIR), Susceptible-Exposed-Infectious (SEI), or Susceptible-Infectious (SI) type. Each strand is artificially seeded into the system at chosen times and can then spread between phones of users. At any given time, a phone can be infected with many strands, and the phone reports its strand infections to the server periodically. Individuals’ identities and social contacts are not recorded in this reporting, ensuring anonymity. A key aim of the Safe Blues idea is to give policymakers another tool that they can use in their effort to visualize the real-time spread of an epidemic. In contrast to those systems that model population contact and implement agent-based simulations, Safe Blues is an emulation of a group of epidemics based upon a contact process that takes place in the population itself.

We devised a campus-wide experiment at The University of Auckland City Campus. This is the first attempt to implement such a system. An outcome of this experiment is an open-source (virtual) epidemic spread dataset which can be used for further modeling, training, and analysis. Our initial plan was to conclude the experiment during November 2021, with the release of data afterwards. However, due to an extensive lockdown in Auckland, the experiment will now run through the second half of 2022. In this paper, our primary focus is on the experiment’s methods and the experience gained. Also, we illustrate general outcomes and results to date. The details we present may be valuable to other teams planning similar experiments in the future. Table 1 describes the phases of the experiment, their timelines, and the period at the University of Auckland during which these phases run.

**Table 1.** The timeline of the experiment at The University of Auckland.

| Phase | Dates | Study period | Comments |
| --- | --- | --- | --- |
| 1 | May 1 - Jun. 10, 2021 | S1, 2021 | Used for debugging |
| 2 | Jul. 18 - Sep. 16, 2021 | S2, 2021 | Lockdown on Aug. 17, 2021 |
| 3 | Sep. 17 - Nov. 4, 2021 | S2, 2021 | Cancelled due to lockdown |
| 4 | Jun. 10 - Sep. 8, 2022 | S2, 2022 | Currently planned |
| 5 | Sep. 9 - Nov. 3, 2022 | S2, 2022 | Currently planned |

As an illustration of the experiment and some of the collected results, consider Fig 1 where we depict the timeline July 28 – September 9, 2021. Phones of participants were “infected” with strands on July 29 and the figure presents the trajectories of the ensuing epidemics along with the number of participants who attended the campus during that period. There are multiple Safe Blues strand trajectories, the (artificial) infection on July 29 included multiple repeats of the same type of strand and multiple types of strands. In fact, not displayed in this figure, about 600 strands were seeded into the participating population. The black trajectory depicts the daily count of campus participants. The weekly attendance pattern, with lower attendance at weekends, can be seen clearly. The green and red trajectories represent the hourly counts of participants whose phones were in the states of *exposed* (infected but not infectious) and *infected*, respectively. As is apparent from the plot, Safe Blues infections continued until the week of August 17 at which point the campus was closed due to a (real) government lockdown. At that point, the number of participants who attended the campus immediately dropped to fewer than 5 per day. As a result, the number of new infections (exposed participants) immediately decreased and within several weeks the number of infected participants also decreased to zero.

**Fig 1.**
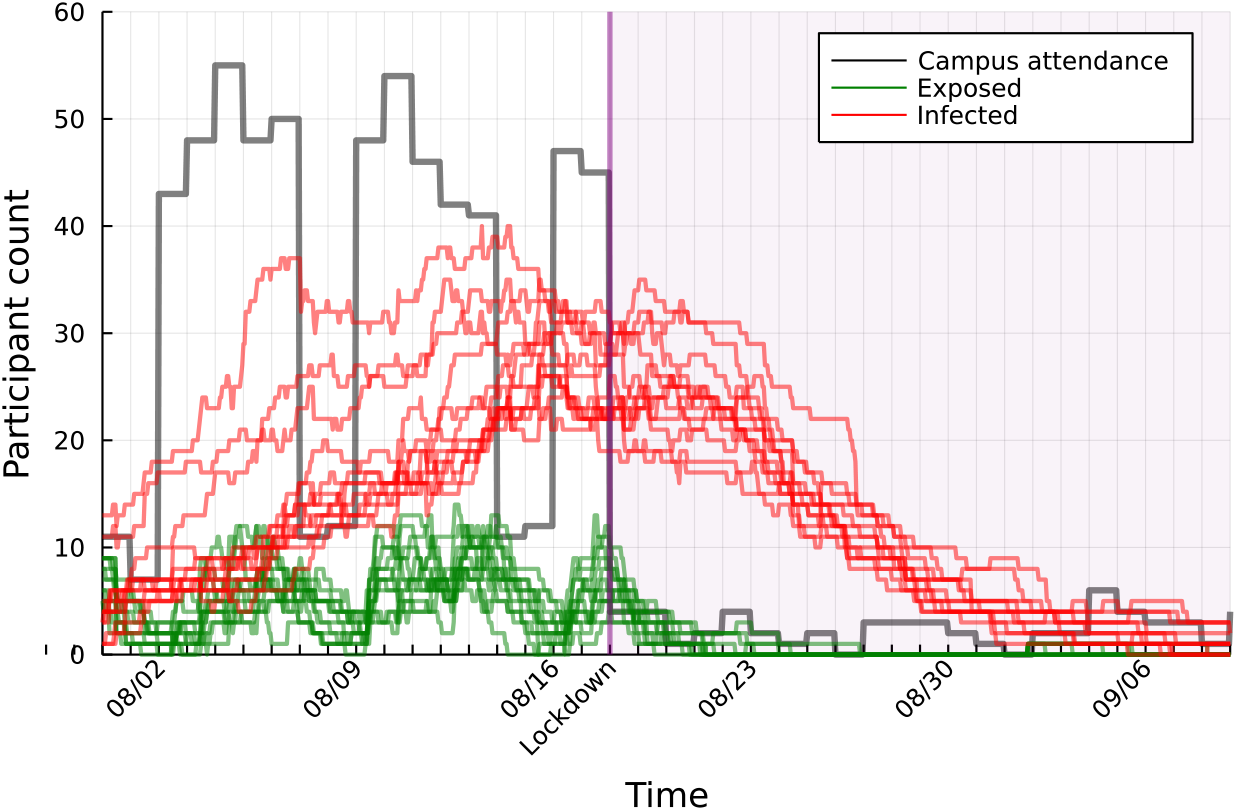
The effect of campus closure, due to actual lockdown in New Zealand on August 17, on the virtual Safe Blues epidemic. Red and green trajectories show the daily counts of participants whose phones were infected (red) and exposed (green) respectively. The black trajectory shows the daily count of participants who attended the campus.

The Safe Blues experiment was not intended to interact with actual COVID numbers or lockdowns. In fact, we chose New Zealand as a destination because it was essentially COVID free for the second half of 2020 and the first half of 2021 and we believed that a university campus could serve as a good first testbed for Safe Blues. In making this decision, we were aware that the university campus did not directly mimic the population dynamics in all of New Zealand. For instance, during the Auckland lockdown in Phase 2, the campus was completely shut down, while in contrast, people in greater New Zealand still interacted, for example, to go shopping. We did not foresee this lockdown in planning the experiment. Nevertheless, the closure of the campus due to the actual physical lockdown served to illustrate the key point of Safe Blues: *safe virtual virus strands that are measured in real-time can give an indication of how actual viruses are spreading, and with enough data and proper machine learning techniques, prediction can take place as well*. The Safe Blues system could thus be applied to predict the spread of viral diseases within a subgroup of the population.

This paper outlines the experimental setup, software, subject recruitment practices, ethical considerations, and dataset description of the experiment. It also presents our current experimental results. Our goal in doing so is to showcase the methodologies and experience gained from the experiment. The source code for the project is freely and openly available at [3]. Further, at the conclusion of the experiment, data will be made available via [4].

## Background

We now present an overview of current practices and specific non-clinical experimental studies that share similar concepts with Safe Blues. Most COVID-19 data are gathered by public health authorities from testing, hospitalizations, and deaths. Various non-government organizations, such as the World Health Organization (WHO) [5], the Center for Systems Science and Engineering at Johns Hopkins University [6], and nCOV2019 [7], collect this data on a global scale and provide daily trend updates. However, such data are prone to lags, biases and inconsistencies, and may not reveal the true characteristics of the disease in real-time. Hence, alternative surveillance methods are needed.

Participatory syndromic-surveillance is one such approach that collects self-reported data on COVID-19 symptoms, test results, and other risk factors for COVID-19 via mobile applications or web-based surveys. Examples of such web surveys include InfluenzaNet [8], FluTracking [9], Outbreaks Near Me [10], CoronaSurveys [11], and the Global COVID Trends and Impact Survey [12]. Among the most recent mobile apps are the COVID Symptom Study [13] and Beat COVID-19 Now [14].

To identify potential COVID-19 hot spots, artificial intelligence is being used in conjunction with information obtained from informal sources, such as Google News, eyewitness reports, social media, and validated official alerts. HealthMap [15, 16], BlueDot [17], and Metabiota [18] are such tools. Early detection of outbreak regions through wastewater examination is also used in some countries [19]. Remote patient monitoring devices, such as continuous wearable sensors (e.g. smartwatches, Fitbit, Oura Ring, WHOOP strap) and smart thermometers, are also being tested as potential tools for tracking COVID-19 [20–25]. These tools measure some of the physiological indicators of an individual’s health, such as temperature, heart rate, blood oxygen level, pulse rate, sleep performance, and step counts, on a daily basis. The device can identify deviations from an individual’s baseline level which may indicate the possibility of an illness developing.

Contact tracing is a popular method for identifying infected but asymptomatic individuals. Under this approach, people who have a history of exposure to a positive case are identified and tested as soon as possible. Various mobile applications and web-based surveys are used for contact tracing [11, 12, 26–33]. Many Bluetooth-based apps, however, were abandoned after their initial release in 2020 [34, 35]. Many jurisdictions have QR code scanning systems in place to track and manage COVID-19. However, data from such apps are typically useless when prevalence increases. For example the Check In Qld app [36] in the Australian state of Queensland was successfully applied to trace contacts of positive cases during 2020 to mid-December 2021 when the Queensland state border was closed and case numbers were single or double digit at most. However, once the border opened and daily new cases grew to three or four digits, the Check In Qld app was generally abandoned.

Moving on to experimental studies, we mention two major non-clinical citizen science experiments conducted prior to the pandemic for disease surveillance purposes. The first is the FluPhone experiment which took place in the United Kingdom between 2009 and 2011 [37]. In this experiment, participants reported their influenza like illness symptoms using the FluPhone app, which also recorded the proximity of participants’ devices via Bluetooth and their location via GPS. The number of people encountered by each participant was then estimated and published on the study website [38]. The FluPhone app, like the Safe Blues app, modeled the spread of virtual SEIR type diseases, allowing participants to see real-time profiles of disease propagation in their contact network [39]. However, unlike the Safe Blues app, which is designed to simulate hundreds or thousands of strands, the FluPhone software was designed specifically to mimic the spread of SARS, flu, and the common cold. FluPhone was a unique experiment at that time, but in contrast to Safe Blues, it was designed with less of a focus on capturing physical social interactions in a privacy preserving manner, and more of a focus on mimicking real disease. The second study is “Contagion! The BBC Four Pandemic experiment”, which also took place in the UK, but this time in 2018-2019. The BBC Pandemic mobile phone app was used in the experiment to record participants’ locations and self-reported contacts. A subset of this dataset was used to simulate various non-pharmaceutical intervention (NPI) strategies, such as case isolation, tracing, contact quarantining, and social distancing, to investigate their effectiveness in limiting the spread of COVID-19 [40].

Since the middle of 2020, many countries have been investigating the risk factors involved in opening their society through mass-gathering experiments. Two well-known examples are the RESTART-19 [41] experiment, which took place in Germany in August 2020, and a study which took place in Spain in December 2020 [42]. Both assessed the risk of COVID transmission during an indoor live concert, using a variety of seating, standing, and hygiene measures, as well as maintaining optimal air ventilation inside the venue. In both studies, contact tracing devices were used to measure contacts made during the event, and PCR tests were performed a few days later. The RESTART-19 study showed that when moderate physical distancing was applied in conjunction with mask-wearing and the conditions for good ventilation were met, indoor mass-gathering events could be held safely. Also, the trial in Spain demonstrated that with comprehensive safety measures, such as face masks and adequate ventilation, indoor mass events could be held without the need for physical distancing.

Some experiments have included a series of mass-gathering events with a variety of indoor and outdoor settings, seated and standing audience styles, structured and unstructured audience styles, and participant numbers. Two such examples are the Fieldlab Events [43] which took place in the Netherlands in February and March 2021, and the Events Research Program [44], which took place in the United Kingdom from April to July 2021. In both experiments, comprehensive public health measures, such as face mask use, hand sanitizing, social distancing, and adequate ventilation at indoor events were observed. Following the events, contact tracing and PCR testing were carried out. According to the Fieldlab Events, outdoor events with 50-75% of the normal visitor capacity could be held provided that strict non-pharmaceutical intervention measures are followed. A robust result from the Events Research Program is yet to be published

Other relevant experiments include a health workers protest [45] in South Korea in August 2020 and a martial arts competition [46] in the UAE in July 2020. During both events, participants were required to wear face masks, practice hand hygiene, and maintain physical distance. COVID-19 symptoms were self-reported by protesters in South Korea after the rally. All PCR tests performed on a subset of rally participants returned negative results. PCR tests were conducted twice weekly during the UAE event, and none of the contestants had positive results, indicating that mass-gathering events with restrictive measures could be held safely.

## Materials and methods

We now describe the experimental setup, ethics, software, participant management, and data collection aspects of the experiment as well as supporting tools such as a simulation model.

### Experimental setup and the Safe Blues system

As stated in the introduction, the overarching purpose of the campus experiment is to test the performance of the Safe Blues system. In doing so, we are interested in assessing the ability to use virtual safe virus-like tokens to predict the spread of pathogens. However, an experiment involving actual biological pathogens, or relying on the actual spread of disease is infeasible and hence our experiment uses measurements from the digital domain. The key question is then to test if the spread of some Safe Blues strands can be detected and predicted by measuring the spread of other strands.

In its most basic form, our purpose is to treat a single strand as a *red strand* which is assumed not to be measurable in real time. Further, we treat all other Safe Blues strands as real-time measurable virtual viruses, namely *blue strands*. The statistical goal is then to benchmark predictions of the future evolution of the red strand based on either,

I. only past measurements of the red strand (a proxy for estimation in the absence of blue strands), or
II. the combination of past measurements of the red strand, past measurements of blue strands, and current measurements of blue strands.

For example, Fig 2, which also appeared in [1], presents a simulation run where blue strands are measured in real time, but the red strand is only measurable with a two-week delay. Here the Safe Blues machine learning framework was used to predict the current unobserved state of the red strand. Similarly, it can be used for near future predictions. However, this figure is taken from Monte Carlo simulations of physical contact processes, and not from actual experimental measurements. The Safe Blues experiment attempts to improve upon this by using the actual physical mobility of individuals. The experiment aims to test whether (II) can yield much better predictions than (I).

**Fig 2.**
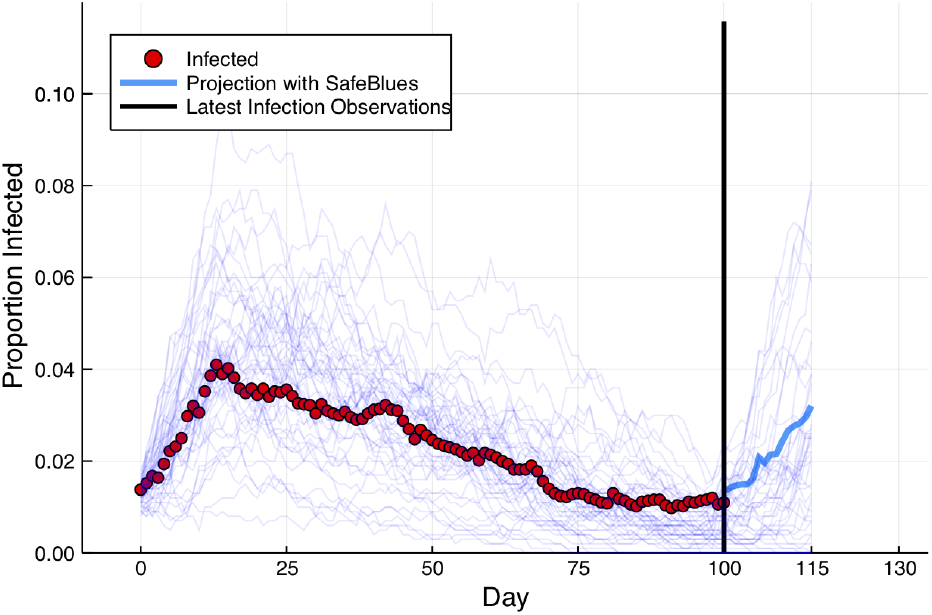
Estimation via simulated epidemics. At day 115, we only have red strand information up to day 100. Nevertheless, current blue strand measurements allow us to estimate the current state of the epidemic during days 101-115.

An additional salient feature of Safe Blues is the interaction with social distancing measures. For this, we would ideally like to ask participants to group together or stay apart similarly to the way that government social distancing measures work. However, this is clearly not feasible with real-life participants and hence the experiment creates *virtual social distancing* to mimic social-distancing measures. The details of how this is done are described below in the subsection Virtual social distancing.

As a first attempt for such an experiment, we chose the University of Auckland City Campus due to the fact that the campus was open to students and staff during 2021 (up until the unexpected lockdown of Aug 17, 2021). The experiment consists of 5 phases. Table 1 provides the timeline of the experiment including the time period of the year, the study period and a brief description of each phase. The target population of the experiment is the student body, but participation is open to any regular attendee or visitor of the UoA City Campus who is at least 16 years of age and uses an Android mobile phone. All participation is voluntary, and at any time, participants could opt-out of the experiment and uninstall the Safe Blues app. By default, participants are invited to join prize draws which we carefully designed to maximize participation (see details in subsection Participant management and ethical considerations below). However, participants are allowed to take part in the experiment without joining the prize draws.

The Safe Blues system is made out of four components: (1) the Safe Blues app, (2) the campus simulation dashboard, (3) the campus experiment leader dashboard, and (4) the Safe Blues data dashboard. All four components are available online at [47], [4], [48], and [49] respectively. Fig 3 displays a snapshot of these four components.

**Fig 3.**
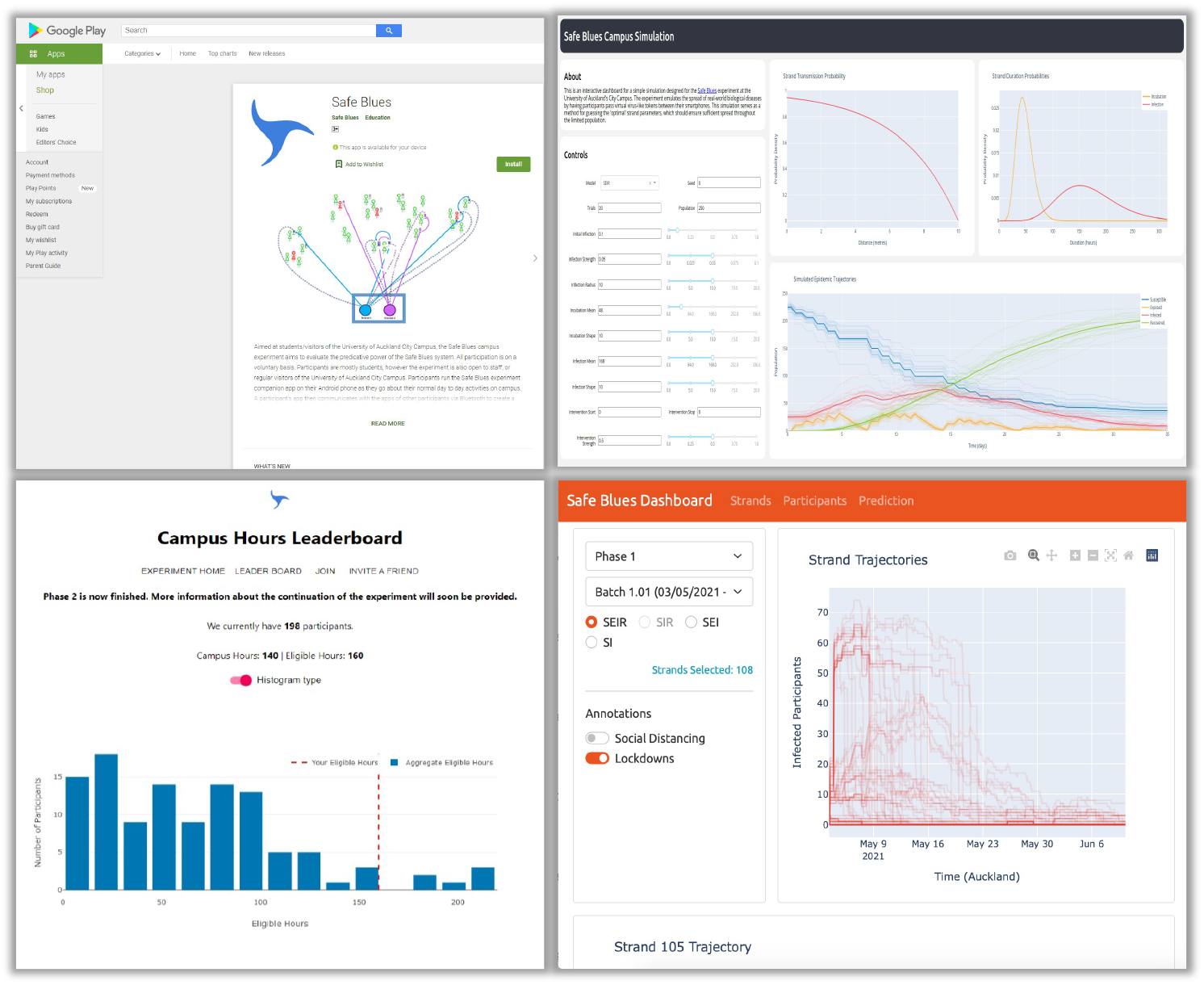
The Safe Blues system: the Safe Blues app (top left), the simulation dashboard (top right), the campus experiment (campus hours leader board on bottom left), and the data dashboard (bottom right).

### Strand and device management

Participants run the Safe Blues app [47] (see also Fig 3 (top left) for an illustration of the app) as they go about their normal day to day activities on campus while enabling Bluetooth and location services. Location services are only needed for prize based rewards as described below. A participant’s app then communicates with the apps of other participants via Bluetooth to pass on digital ‘virus-like’ tokens, namely Safe Blues ‘strands’. This simulates an epidemic spreading through the community. There are many types of strands of such virtual safe epidemics, and the live emulation of all of the epidemics happens in parallel driven by the actual physical contact processes of participants.

The app is not malicious and does not interact with any other app that users may be running on their phone. Open-source code is available on GitHub via the Safe Blues website [50]. Nevertheless, the app, like any other mobile app, consumes the battery of the phone. It is the participants’ responsibility to manage their phone battery usage, and our experience has shown that some participants turn off the app while away from the campus. The app is only available for Android due to the fact that iOS phones cannot run such an app in the background. This clearly limits the participating population. We discuss the technical details of the app software in S2 Appendix.

With the exception of participant reward information, described in subsection Participant management and ethical considerations below, information recorded by the Safe Blues system is limited to the aggregated counts of each strand. Every 15 minutes a phone uploads the status of its infections in terms of ‘exposed’, ‘infected’, and ‘recovered’ for each strand. The total number of ‘susceptibles’ is then inferred based on the total number of phones participating at any given time. This uploading occurs via a temporary anonymous 256 bit ID which changes every 24 hours on the phone. Thus the Safe Blues server does not keep track of the individual infections of phones and it cannot uniquely identify a phone beyond a 24 hour period. The temporary ID is still useful for correct counting of infections on the server side, since messages are sometimes lost or not sent if the phone is without connectivity (see S4 Appendix where we describe the algorithm for interpolation and imputation of counts to handle this). The individual phone strand information is never cross-referenced with private participant information, further preserving the anonymity of participants.

The injection of new infections into the participating phone population is carried out via an API (Application Program Interface) available to the phones. In each phase of the experiment we inject multiple strands with each batch containing a collection of individual strands. For example, in phase 1, where we focused on testing and tweaking the system, there were 7 batches in total labeled `1.01` to `1.07`. Similarly, in phase 3 there were 22 batches in total, labeled `3.01` to `3.22`. We discuss the number of strands in each batch, their parameters, and their purposes in the experiment in S5 Appendix. When a batch of strands is ‘injected’, all participating phones become aware of the strands of the batch and each strand has a pre-specified seeding probability which is typically set to 0.05, 0.1, or 0.2. Then at a specified start time, each phone is independently infected by the new strand in accordance with the seeding probability. This ‘seeding’ of new strands thus emulates the arrival of new outbreaks of the epidemic into the population.

We applied four types of epidemic models for the phone population: SEIR, SIR, SEI, and SI. In both SEIR and SEI models, when a susceptible phone receives a strand, it first becomes exposed and remains in this state for a random time period known as the incubation period. During the incubation period, the phone is unable to infect other phones. After the incubation period, the phone becomes infected and remains in this state for a random time period known as the infection period. During this time, the phone has the ability to infect nearby phones by exchanging a Bluetooth token. The distribution of the incubation and infection periods is explained further below. The infection period in the SEIR type epidemic is finite, and the phone stops infecting other phones at the end of this period. Consequently, its state is labeled as recovered. The infection period in the SEI epidemic is infinite, and the phone cannot recover. There is no incubation period in the SIR and SI epidemics, and a susceptible phone becomes infected immediately after receiving the strand. The SIR epidemic has a finite infection period and the phone recovers at the end of it. The infection period of the SI epidemic is infinite, and the phone remains in the infected state throughout the epidemic.

During phases 1-3, and including the intervening period between phase 3 and phase 4, we injected 4155 strands in total into the system. Of these, 28% are of the SI or SEI type (not involving removal/recovery) and the remaining 72% are of the SIR or SEIR type (allowing recovery). Fig 4 depicts the cumulative counts of strands released over time during phases 1-3.

**Fig 4.**
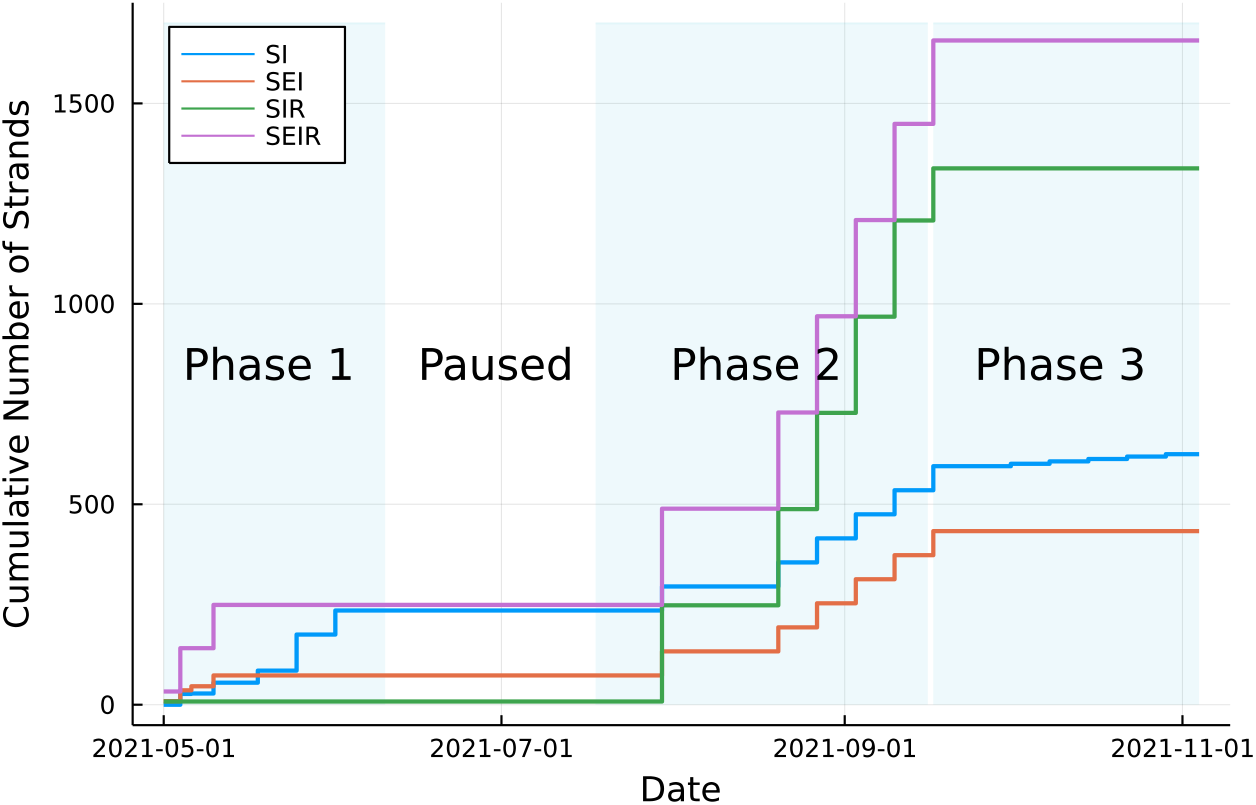
The cumulative number of strands over time broken up into SI, SEI, SIR, and SIER types and phases of the experiment.

Beyond the classification of SI, SEI, SIR, and SEIR, each strand, uniquely identified by a `strand_id`, which has specific parameters that influence its spread. A full specification of the protocol for these parameters is in Appendix A1 of [2]. However, the protocol there does not deal with specific distributional information and the infection probability mechanism. Hence we now outline these details.

We use gamma distributions for both the incubation and infection times and parameterize them by a mean *μ* and a shape parameter *k*. That is, for each *x* ∈ (0, ∞), the probability density function is

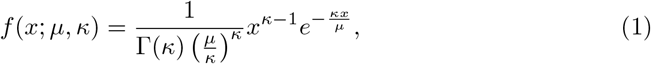

where Γ(·) is the gamma function. In this case, the ratio between the variance and the square of the mean is 1*/k*. In many cases we used *k* = 3; in other cases to create (nearly) deterministic times we set *k* = 10, 000.

The other important strand information deals with the probability of infections of nearby phones. At every time where two participating phones are near each other, Bluetooth messages are exchanged during a *session* and throughout this session, distance measurements are carried out. In principle using individual Bluetooth messages may appear to be preferable to creating sessions. However the nature of the Bluetooth protocol and the underlying software implies that sessions are the preferable technique; see S2 Appendix.

At the end of the session (capped at 30 minutes), the median distance from all messages is computed and denoted by *d*. The duration of the session is denoted by *t*. We expect that with closer distances and longer duration, infection is more likely. We chose the probability of infection parameterized by the strand’s strength, *σ*, and the maximal infection distance, *ρ*, to be

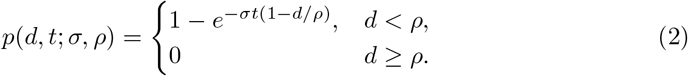

In general, we expect strands with higher *ρ* or higher *σ* to be more infectious.

In setting the strand parameters *σ* and *ρ* we initially used a simulation model (see subsection A campus simulation model for details). Subsequently, we adjusted the parameters based on field experience (see section Results and discussion section for details).

### Participant management and ethical considerations

Our goal in participant management is to motivate participants to run the Safe Blues app while on campus. A first decision was whether to couple participation data with strand data (number of virtual infections). We chose not to do so. While such coupled data could be useful, our primary goal is the strand-count time series for which coupling is not needed.

A second decision was whether to pay participants a ‘flat rate’ for participation, for example with coffee vouchers proportional to their participation hours or to use prizes. As the total budget was limited, and in accordance with other experimental research [51, 52], we opted for prizes. As this is a digital experiment we chose iPad, Android phone, and Fitbit prizes, with 9 prizes per prize draw. See S1 Appendix for details of the prize draw rules.

Participants were recruited directly via online flyers, posters and videos. To take part in the experiment, participants first needed to install the Safe Blues Android app on their mobile phones. This gave them a random 10 digit ID which identifies them only for purposes of experiment participation and prizes but is not associated with their strand infections. With this ID, participants can then register their email address which is used for communicating experiment messages and prize winners.

As participants enter the city campus, an Android geofencing mechanism spawns an event on the app, and then when they leave the campus (leave the geofence) an additional event is spawned. A message of participation hours is recorded on a server. The participation hours contribute to the chance of winning a prize. In general, the more hours a participant runs the app on campus, the higher the chance of winning a prize (see S1 Appendix). The app does not track the location of participants with the exception of indicating whether or not the participant is within the campus geofence area. Fig 5 (left) displays a snapshot map of the UoA City Campus with the geofenced area marked on it.

**Fig 5.**
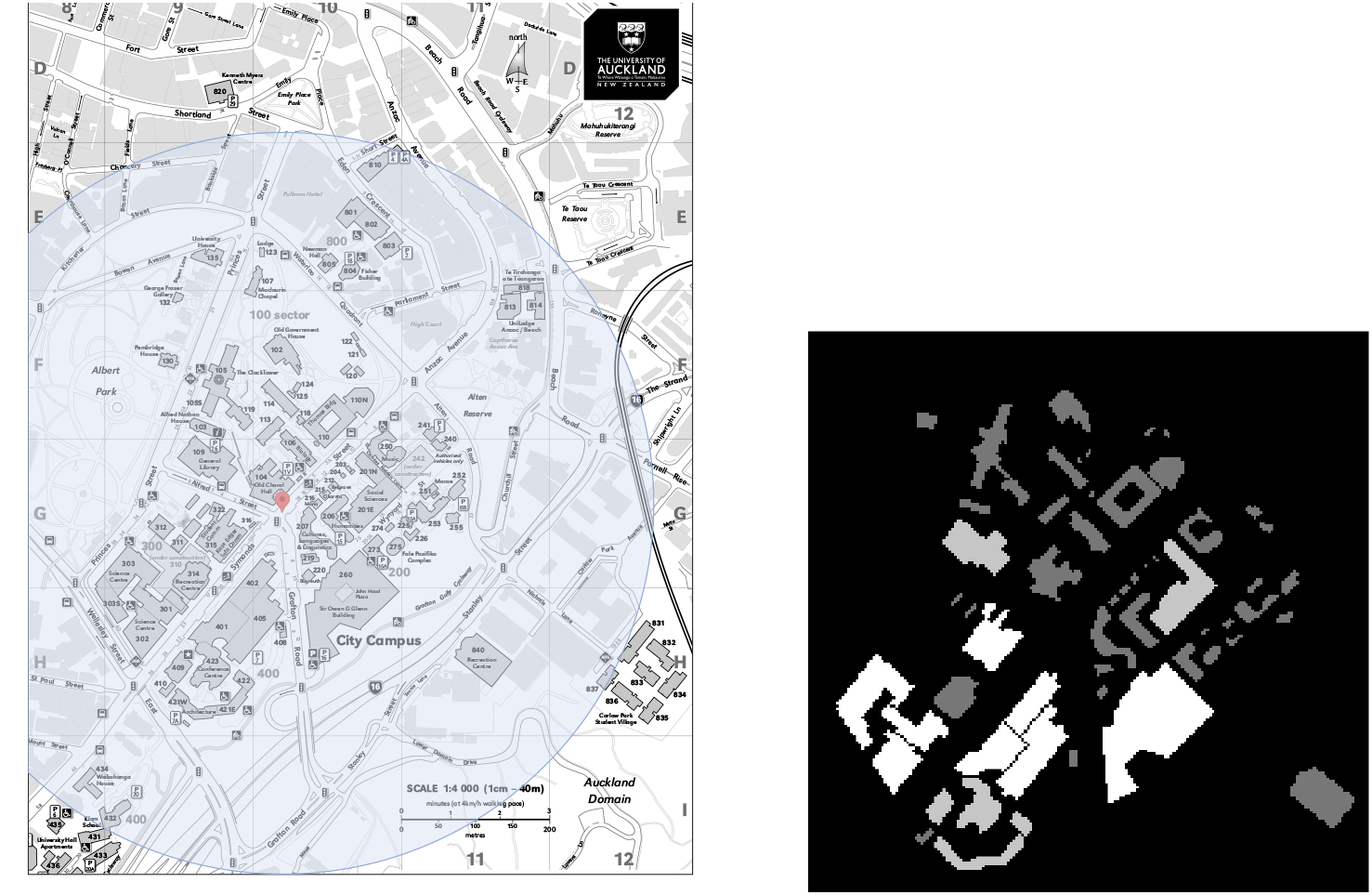
The University of Auckland City campus with the geofenced area supporting the experiment marked as a circle (left). A heatmap representation of buildings used in the simulation prior to the experiment (right).

As an additional side-benefit of the experiment, we provide the aggregated visits to campus and duration statistics as part of the dataset. The left side plot in Fig 6 depicts the daily number of participants who were registered, reporting, and attending the campus in the experiment during phases 1–3. The right side plot in this figure displays the distribution of the means for the daily campus hours collected by participants, over weekdays and weekends, during phases 1–3. As per the prize draw rules (S1 Appendix) the maximum daily campus hours that each participant can collect is capped at 10.

**Fig 6.**
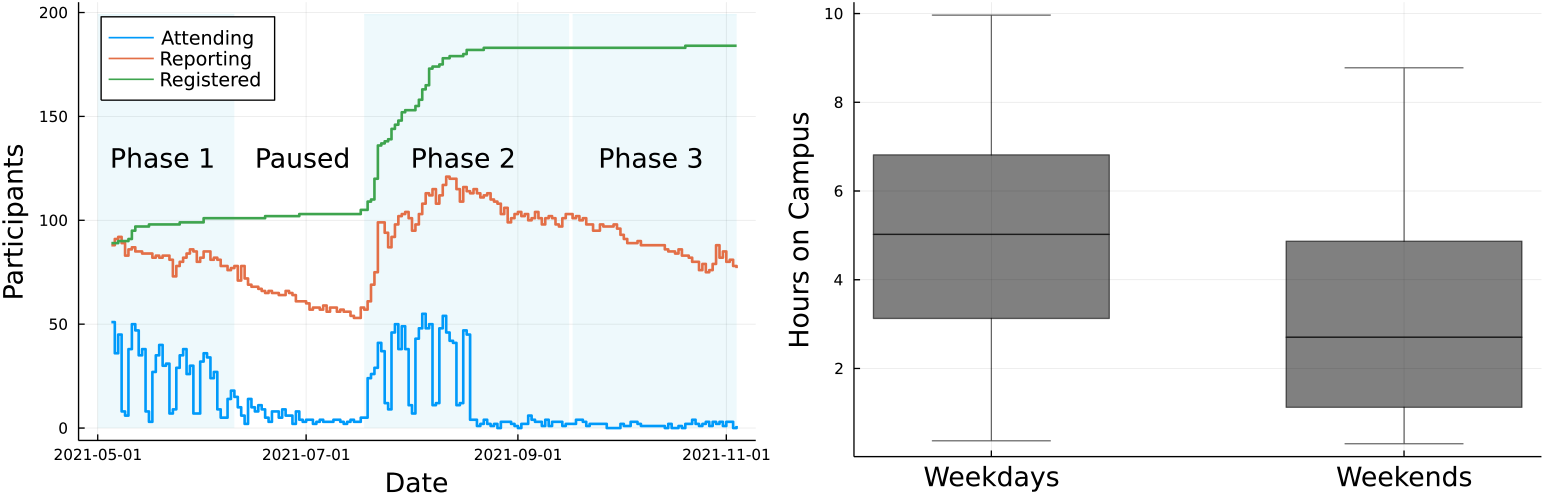
Evolution of the daily number of participants who were on campus (Attending), reporting, and the cumulative number of participants who were registered in the experiment, during phases 1 to 3 (left). Box plot of the means from the 5 number summaries for daily campus hours (right).

By the end of phase 2, about 20% of the registered participants were not running the app. In an attempt to enroll more participants in the experiment, we upgraded the reward scheme during phase 2. This included an ‘invite-a-friend’ option, which increased a participant’s chance to win a prize if new participants joined the experiment through their invitation. Those who joined the experiment through the invite-a-friend mechanism were also rewarded with bonus eligible hours. See S1 Appendix for further details about rules and the invite-a-friend mechanism. Joining the prize draws was not compulsory for those taking part in the experiment.

Although we were not conducting any clinical or health research including human data, we required approval from the University of Auckland Human Participants Ethical Committee (UAHPEC) before doing any form of research involving university volunteers. The study was approved by UAHPEC under ethics number 22143 in March 2021. In this application we addressed ethics considerations, including naming all researchers, description of the study, location of study, methodology, participants and recruitment process, data management, funding, Māori-focused consultation and engagement, and consistency with the principles of Te Tiriti o Waitangi.

We also provided the ethics committee with a copy of all the Safe Blues website pages, a permission letter from course directors (for big statistics courses where the project was advertised) and head of the Department of Statistics, participant information sheet, poster, the consent form, the data management plan, and the observation schedule.

### Data management

The experiment is managed through two distinct databases. A Participant Management System (PMS) is used to store the email addresses and consent agreements, as well as a record of the campus hours. The PMS is hosted at the UoA, and data are used only for the purposes of managing the prize draws and the list of participants. Data in the PMS is completely disconnected from the experiment data and will not be publicized in any way, with the exception of analysis of aggregated participation counts over time (for example, see Fig 6).

The second database, called the Anonymous Data Server (ADS), is managed in the cloud and contains an aggregate, anonymized, time-stamped record of the number of phones with each strand. For each strand, we record this data on an hourly and daily basis, and indicate the aggregate number of phones in each epidemiological state (susceptible, exposed, infected, recovered) over time. As this database follows the Safe Blues protocol, phone (app) identities are not revealed during communication, and phones (apps) only have temporary IDs that are replaced on a daily basis. We spell out the technical details of how we record data in the PMS and ADS in S3 Appendix.

We record aggregated participation counts from the PMS as daily and hourly measurements, along with the daily and hourly means and five number summaries–that is, the minimum, first quartile, second quartile, third quartile, and maximum–of campus hours in a CSV file. On days where there are fewer than 5 participants these numbers are omitted for privacy reasons. Similarly, the aggregate Safe Blues data from the ADS are also stored in several CSV files, one for each strand. See S6 Appendix for specific details of the Safe Blues data repository. The Safe Blues data will be made publicly available in the Safe Blues data repository after the experiment concludes. Plots of the data are currently available as a web-based dashboard at [49]. See Fig 3, bottom right, for a snapshot of the dashboard.

By agreeing to take part in the experiment, a participant agrees to share the Safe Blues data of their Safe Blues app. At any point in time, a participant may choose to withdraw from the experiment and this will result in deletion of their personal information from the PMS. However, their aggregated anonymized data already recorded on the ADS will remain in the database and will potentially contribute to the scientific findings of the experiment.

### A campus simulation model

We used a simple simulation model to approximately capture the expected behavior of the participants, and to aid with our initial choice of strand parameters to be used in the experiment. This discrete-time stochastic spatial compartmental SEIR model was used as an initial guide for ranges of the maximal infection distance and infection strength parameters in Eq (2).

A Safe Blues strand is characterized by its seeding probability, *π*, infection strength, *σ*, maximal infection distance, *ρ*, incubation time distribution, and infection time distribution. In the simulation, the initial infections were determined by Bernoulli random variables with each simulated participant independently having a chance *π* of becoming infected when the strand was activated. The remaining participants could only become infected by being a ‘close contact’ of an already infectious person. After each time step, the positions of participants were independently drawn from a heat map designed to resemble likely locations attended by participants in the real-world experiment; see Fig 5 (right). The time step considered here was 1 hour, which corresponds to the duration of a lecture. Each susceptible individual who was *d* meters away from an infectious individual for *t* minutes was infected with the probability given by Eq (2).

After a strand was successfully transmitted to a susceptible individual, they became exposed and remained in this compartment for a random amount of time (incubation time) drawn from a gamma distribution with mean *μ*_*E*_ and shape *κ*_*E*_ as in Eq (1). Subsequently, once their exposure time elapsed, they became infected and were able to infect further individuals with this strand. The duration of their infection (infection time) was again gamma distributed with mean *μ*_*I*_ and shape *κ*_*I*_. A strand without incubation (SI or SIR) can be described by setting *μ*_*E*_ ≈ 0 and 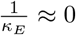 and a strand without recovery (SI or SEI) can be described by setting 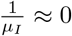 and 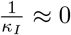. Further details are in the code repository within the Safe Blues GitHub repository [3].

We developed a web-based interactive simulation dashboard; see Fig 3 (top right) and [48]. The interactive dashboard has a variety of control features that enable a user to set the model and its parameters. We used the simulation to focus on exploring parameters for a strand’s infection strength *σ* and maximal infection distance *ρ*. In particular, we explored *σ* in the range [0, 0.1] and *ρ* in the range [0, 20], and fixed the remaining parameters at a specific values. We chose *π* = 0.1, *μ*_*E*_ = 24 hours (or a single day), *κ*_*E*_ = 5, *μ*_*I*_ = 168 hours (or a single week), *κ*_*I*_ = 5.

The simulation indicated that with a population of 50 or more participants, some level of Safe Blues spread is possible. Further we simulated epidemics on population sizes 100, 200, and 500. Based on 1000 simulation runs of the model, we observed that,

1. there existed a minimum value for *ρ* for sustained transmission, which decreased as population size increased, and
2. there existed a region of transitional parameters, which narrowed as population size increased.

These observations allowed us to conclude that *σ* ∈ [0, 0.05] and *ρ* ∈ [10, 20] were the ideal parameter ranges for a population size of 100, *σ* ∈ [0, 0.05] and *ρ* ∈ [5, 15] the ideal parameter ranges for a population size of 200, and *σ* ∈ [0, 0.04] and *ρ* ∈ [2, 12] the optimal parameter ranges for a population size of 500. These observations then guided our initial strand parameter choices.

### Virtual social distancing

Apart from using Safe Blues as a tool for collecting data on virtual epidemic spread, we also tested it as a means to explore how ‘virtual social distancing’ affects these epidemics. Our goal in implementing ‘virtual social distancing’ was to provide a rich dataset which could be used by researchers or public health bodies to explore future intervention strategies through social distancing.

In order to implement the ‘virtual social distancing’, we tweaked the measured (observed) distance, *d*, in Eq (2) by a ‘social distancing factor’. Thus, if the measured distance was 4m and the social distancing factor was 1.5, then the distance, *d*, for infection computation was 4 * 1.5 = 6m. See the Results and discussion section below for results of this testing mechanism.

## Results and discussion

### Calibration of the maximal infection distance parameter

At the start of the experiment we used the parameter ranges determined from the campus simulation study as an initial guide for choosing strand parameters, *σ* and *ρ* as in (2). Our initial purpose was to find dynamic ranges of both the infection strength, *σ*, and maximal infection distance, *ρ*, that affect the spread of strands. Initial results from the campus experiment immediately confirmed the effect of the maximal infection distance, while the effect of infection strength was not apparent in the dynamic range of values that we used. We then further explored the range for the maximal infection distance parameter using the strands released in batches `1.05` and `1.06`, during phase 1 trials. In particular, we experimented with the maximal infection distance parameter within the range [7.5, 500] while choosing the strength parameter within the range [0.1, .24].

In batch `1.05`, we released 30 SI type strands with the strength parameter fixed at 0.16 for all strands and the maximal infection distance parameter chosen from the set {7.5, 15, 30, 60, 120, 500}. We observed that, in general, there were 3 ranges for the distance parameter that produced distinct epidemics. Specifically, the epidemics established when the maximal infection distance parameter was greater than 30. Increasing the maximal distance parameter beyond 120 did not necessarily produce more severe epidemics. Further, the epidemics did not propagate for most of the strands when the maximal distance parameter was less than 30. With this observation in mind, we fine tuned the search grid for the maximal distance parameter using the strands released in batch `1.06`. We released 90 SI type strands in that batch, with the maximal distance parameter chosen from the set {20, 26, 34, 44, 57, 74, 97, 125, 163, 212} and the strength parameter chosen from the set {0.1, 0.16, 0.24}.

The plots in Fig 7 depict the effect of varying the maximal infection distance parameter on the propagation of epidemics. The left side plot shows the daily infection trends of strands in batch `1.06` over 5 days categorized by 3 maximal infection distance ranges. The right side plot displays the difference in infection count between the 1st day and 5th day for the strands in both batches as the distance parameter varies. In both these plots we ignored the effect of infection strength since its confounding effect was negligible.

**Fig 7.**
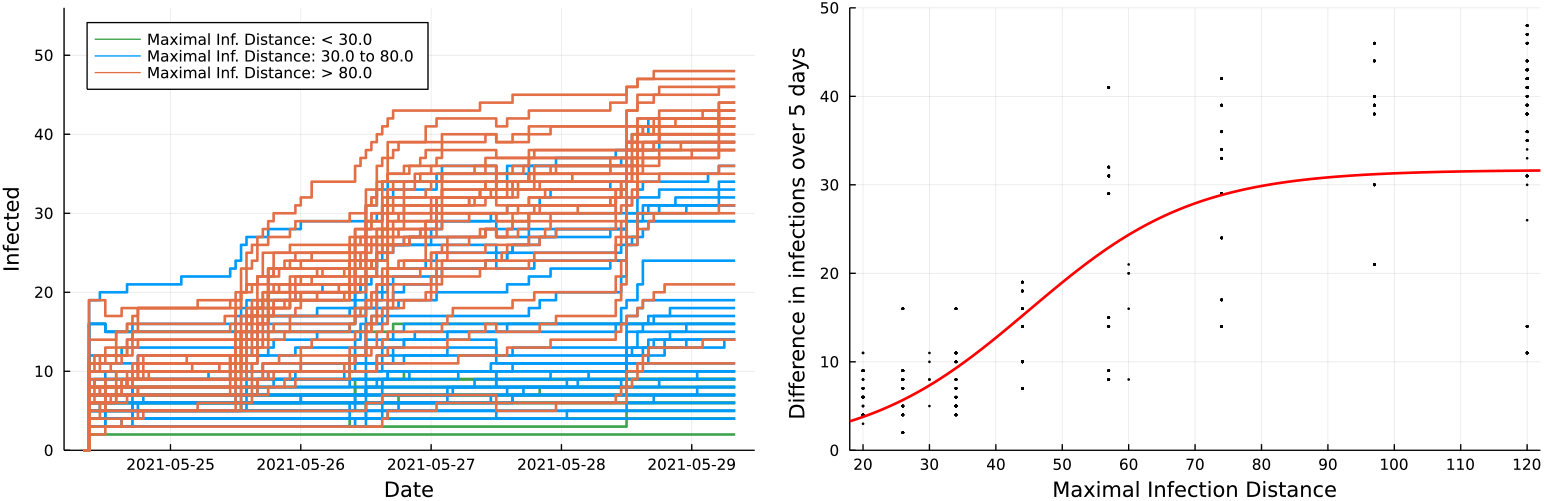
Infection trends over 5 days for strands in batch `1.06`, categorized by 3 maximal infection distance ranges (left). Effect of varying maximal infection distance on the difference of infection (day 5 - day 1) for strands in batches `1.05` and `1.06` (right). The red curve on the right side plot is a fitted sigmoid function.

We fitted a two parameter scaled, and shifted sigmoidal curve to the data, plotted in red. The curve clearly indicates an upward infection effect of the maximal infection distance with saturation at distances over 80 meters, and less than 30 meters. This is not unexpected based on our design of the infection formula (2), yet we initially found the magnitude of the distances puzzling. One may expect Bluetooth transmission to be effective at distances that are significantly shorter. Towards that end, we believe the observed distance, *d* in (2), may be skewed in our app measurements which are based on averaging of RSSI Bluetooth signal strengths. Such bias between the actual distance of devices and the observed distance, *d*, may be further investigated via direct phone to phone measurements of the app. We have yet to carry out such measurements to completion, but initial tests indicated of a mismatch of the order of 30 meters, meaning that phones that are *x* meters apart perceive a distance in the order of *d* = *x* + 30.

### Herd immunity

Herd immunity occurs when a significant proportion of a population become immune to an infectious disease through either vaccination or previous infection, making the disease unlikely to spread within the population. The ‘herd immunity threshold’ is the minimum proportion of the population that must be immune in order to achieve herd immunity. In the simplest SIR model, this quantity can be calculated as 1 − 1*/R*_0_, where *R*_0_ is the basic reproduction number of the disease [53]. The basic reproduction number is the expected number of secondary infections caused by a single infectious person in an otherwise susceptible population [54, 55]. The basic reproduction number for the delta variant of COVID-19 is estimated as 5.1 [56], and thus the herd immunity threshold for the delta variant is approximately 80%.

In our experiment, we observed the herd immunity phenomenon from some of the strand’s epidemics. For example, Fig 8 displays the epidemics of two SEIR type Safe Blues strands with different mean infection periods. It is evident from the two plots that about 80% of the participating population recovered and about 20% remained susceptible after the disease died off, depicting the herd immunity phenomenon.

**Fig 8.**
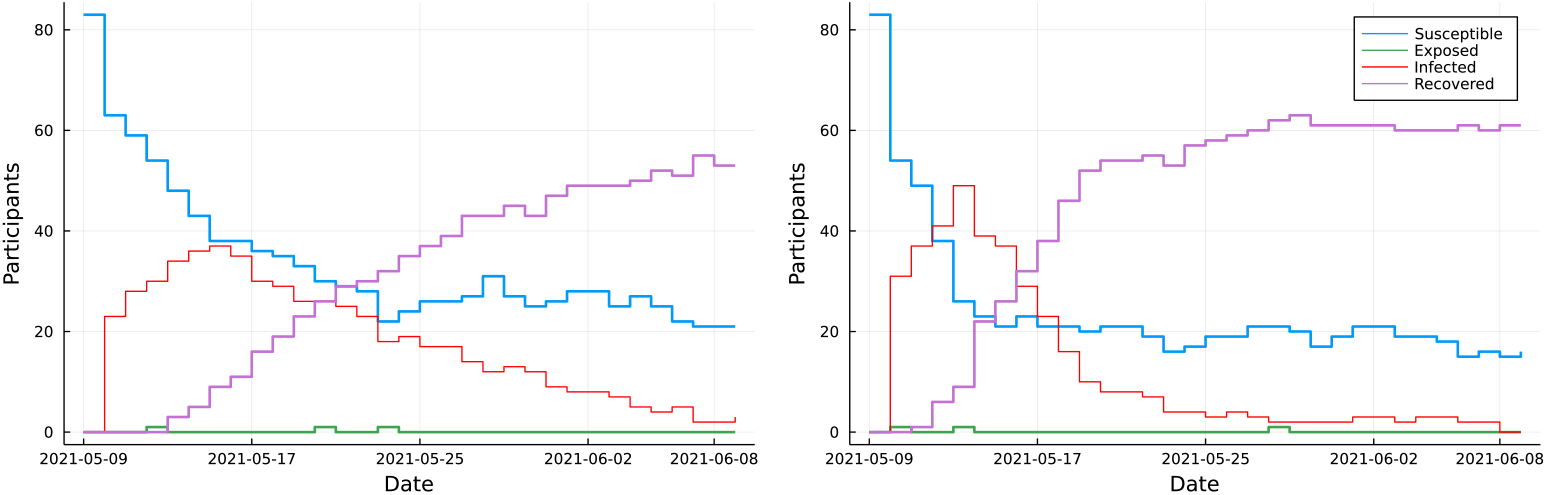
Epidemic trajectories of an SEIR type Safe Blues strand with mean infection period 10 days (left), and 5 days (right). The remaining strand parameters are common to both strands.

### Testing the virtual social distancing mechanism

Social distancing is intended to increase spatial separation. It could be implemented by putting a lower bound on *d* or scaling *d* up. As mentioned previously, we implemented virtual social distancing by multiplying the observed distance parameter, *d*, in Eq 2 by a given social distancing factor. We considered 4 social distancing factors; 1 (no social distancing) 1.25 (low), 1.5 (medium), and 3.0 (high). We tested virtual social distancing for the strands released in batch 1.07, starting from the 3rd day of their release. This batch comprised 60 SI type strands.

We compared each strand’s infection counts on the days prior to and after virtual social distancing was imposed. Fig 9 displays the effect of social distancing and the maximal infection distance on infection counts of three strands. Each data point is comprised of number of infections one day before the virtual lockdown and number of infections one day after the virtual lockdown. We saw three distinct patterns for the counts one day prior to implementing virtual social distancing. When maximum distance was 40, the counts were less than 2, when it was 60, the counts were between 2 and 6, and when it was above 80, the counts had similar values. This distinction was not visible for the infection counts after imposing virtual social distancing.

**Fig 9.**
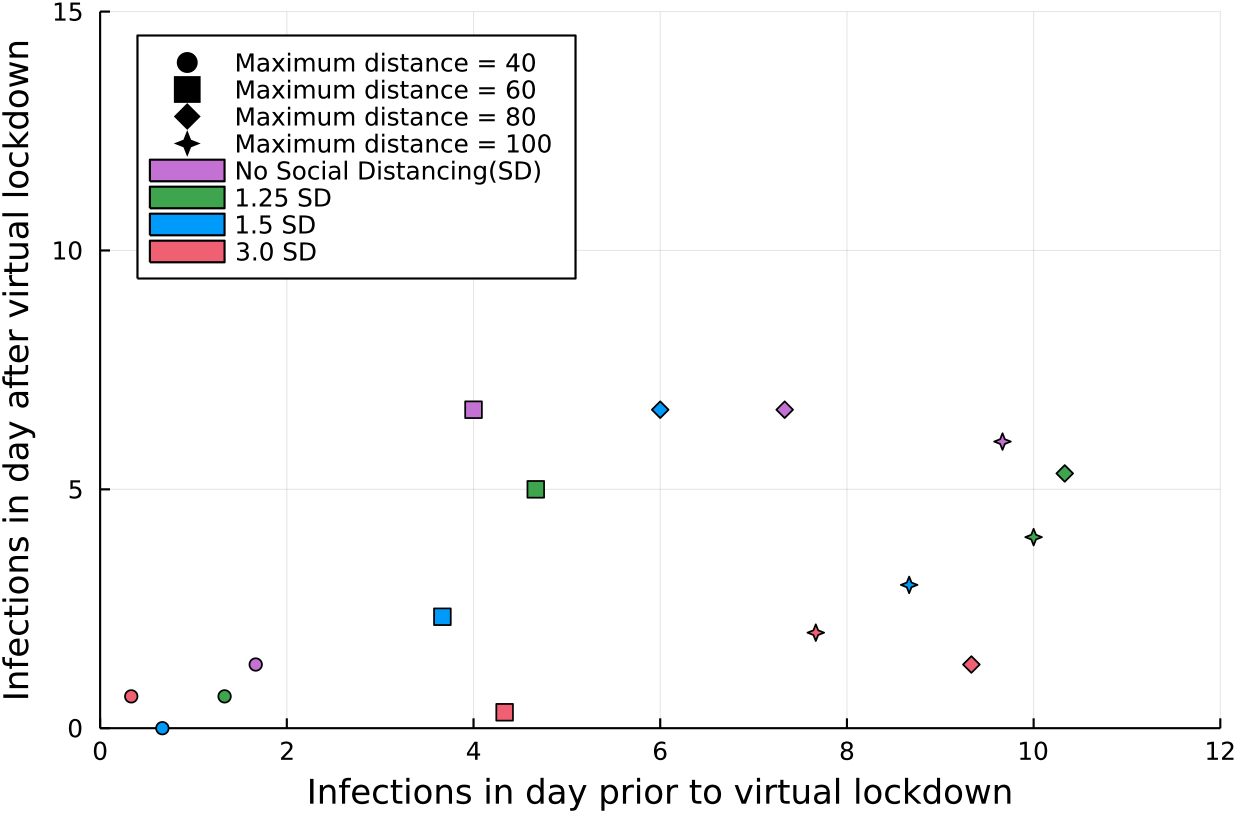
Infection counts of three Safe blues strands on the day prior to and after implementing virtual social distancing, categorized by their maximal infection distance. Each point is the centroid of the triangle formed from the infection counts of the three strands. Social Distancing (SD) is categorized as; 1.0 (no SD), 1.25 (low SD), 1.5 (medium SD), and 3.0 (high SD).

In terms of the effect of the social distance factor, we can observe that infections on the day after the virtual lockdown were, in general, lower for the high (3.0 SD) and medium (1.25 SD) factors. This result highlights the fact that our initial trials on virtual social distancing had an impact in reducing the severity of the epidemics. Higher participant numbers and using more strands would probably strengthen this calculation. We intend to explore virtual social distancing in our future trials once the experiment restarts in 2022.

### The effect of an actual lockdown on the experiment

In the previous section we highlighted that we were able to observe reduction of strand infection counts based on artificially imposed social distancing. However, we were able to observe the same phenomena after the actual lockdown that occurred in New Zealand during phase 2 trial of our experiment.

The actual lockdown in Auckland, took place on August 17, 2021 at the time when we released batch `2.01` strands, and the lockdown was later extended until the end of 2021. There were 600 strands in total in this batch. These were the first set of experimental strands that were released after we determined the maximal infection distance parameter ranges and tested virtual social distancing. The campus was shut down due to the lockdown, and the number of attendees on campus immediately dropped. Consequently, we saw an immediate reduction in the number of exposed participants, and within weeks the number of infected participants reduced to zero. Thus, our data showcased the effectiveness of the actual lockdown.

In Fig 10 we depict the infection trajectories of all strands in batch `2.01`, categorized into their model type (SEIR, SIR, SEI, SI). For all model types we can see a clear effect of the actual lockdown on their strand’s trajectories. As expected, for both the SEIR and SIR models (top two plots in Fig 10), the infection trends gradually reduced or remained close to zero within weeks after the lockdown, and for both the SEI and SI models (bottom two plots in Fig 10), the infection trends stabilized. We also see that the effect of the lockdown was immediate, showcasing the real-time nature of Safe Blues information.

**Fig 10.**
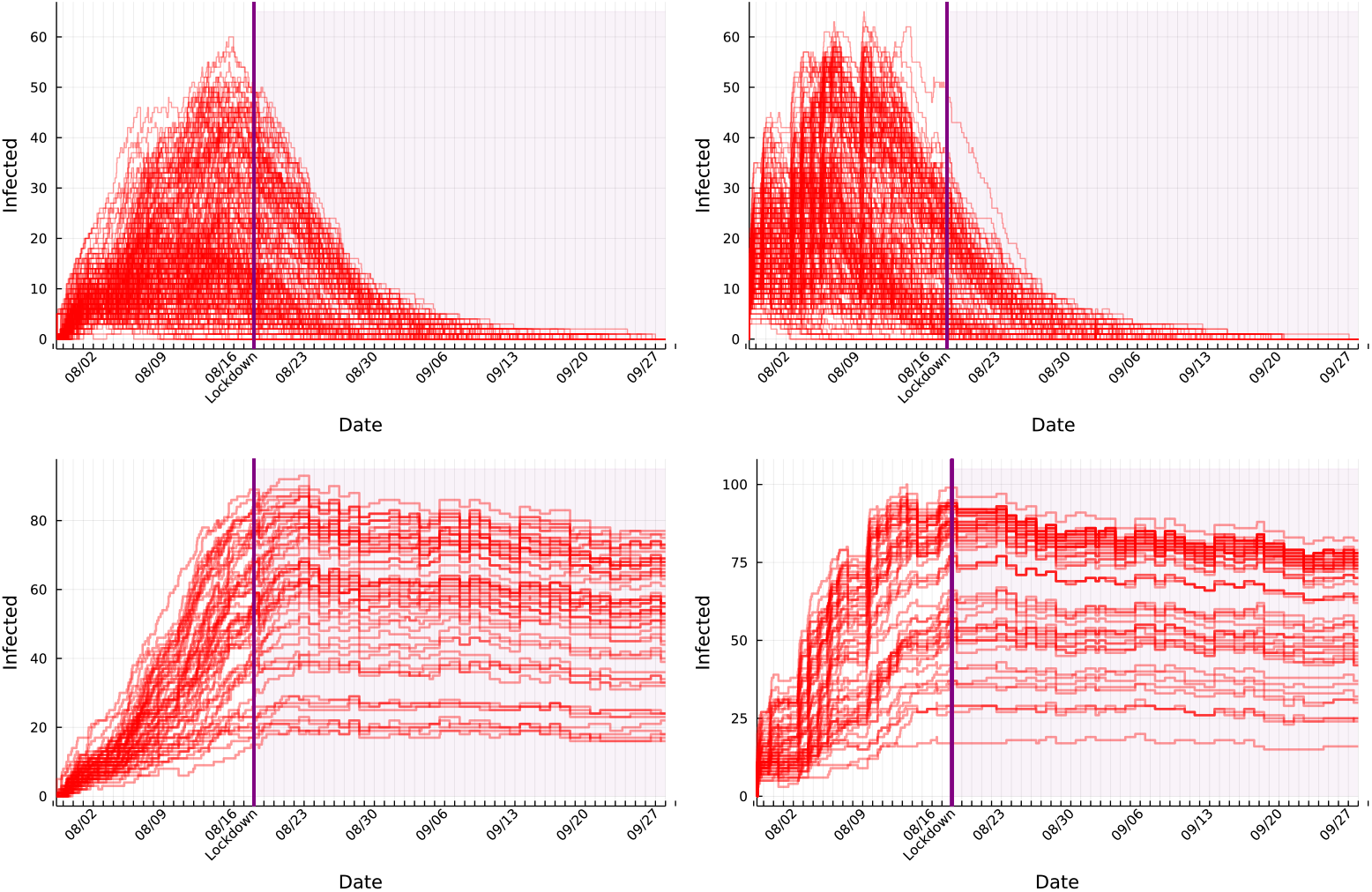
Infection trajectories for all strands in batch 2.01, categorized into SEIR (top left), SIR (top right), SEI (bottom left), and SI (bottom right).

## Conclusion

We have described the design of a Safe Blues experiment in Auckland, New Zealand. The experiment was interrupted by a lockdown in August 2021, so we have extended the experiment to include two more phases once the second semester begins in June 2022. The partial data we collected while calibrating the experiment in early phases, especially after the lockdown, suggests that Safe Blues data will be a valuable tool in the fight against pandemics. In particular, we saw the effect of the lockdown immediately in the strand data, even though in reality the effect of the lockdown would not be observed for several days, since infection counts lag true infections by the incubation period together with the time it takes to get tested and recorded the test result which can be several days. The value of Safe Blues real-time data is even greater in the presence of under-reporting of cases, which arises in the presence of asymptomatic infections, or skepticism or ignorance of the value of reporting or where the efficiency of other methods (contact-tracing, wastewater monitoring or even PCR with group testing) is reduced when prevalence is high.

Our focus in this paper was on the design of the experiment. We discussed the databases, strand management, measures taken to ensure participant privacy, and participation incentives that are essential in such an effort. A simulation tool was useful in initially calibrating strand parameters, but data from initial phases became more valuable than simulation for full calibration. In the early stages of the experiment we were able to approximately model the effect of social-distancing mandates and the results suggest the potential for the full experiment to showcase the potential effectiveness of such measures, as did the data from the true lockdown. As part of the experiment we have built a number of visualizations and these, along with all necessary source code, are available on an open repository. We look forward to providing data from Phases 4 and 5 of the experiment once these are complete.

## Data Availability

At the conclusion of the experiment, data will be made available via https://safeblues.org/data

## Data Availability

At the conclusion of the experiment, data will be made available via https://safeblues.org/data

## Supporting information

**S1 Appendix. Prize draw rules**. Details of Prize draw rules.

**S2 Appendix. App software**. Details of the Safe Blues app software

**S3 Appendix. ADS and PMS servers**. The ADS & PMS technical details

**S4 Appendix. Interpolation and imputation algorithms**. Details of the interpolation and imputation algorithms,

**S5 Appendix. Strand details**. Details of strands and their purposes

**S6 Appendix. Data structure**. Structure of Safe Blues dataset

## Appendix 1: Prize draw rules

As participants run the Safe Blues app within the geofence area (See Fig 5), they collect hours which increase their chance of winning prizes. This is done by accumulating *campus hours* which are then translated into *eligible hours* via the rules described below. During Phase 1 we used a simpler set of rules, and introduced the ‘invite a friend’ mechanism from Phase 2 onward intending to increase participant numbers. See also the prizes web page, https://safeblues.org/prizes/.

### This is an overview of the rules

- For every hour running the app on campus, one *campus hour* is collected.
- Campus hours are accumulated since joining, or since the start of the current phase (campus hours are not collected during *paused periods* (see Table 1).
- A participant may collect up to 200 campus hours per phase.
- A maximum of 10 campus hours per day can be collected.
- Campus hours are converted to *eligible hours* as follows:
  – Each of the first 20 campus hours counts for 2 eligible hours.
  – After the first 20 campus hours, each additional campus hour counts as 1 eligible hour.
- Participants may obtain more eligible hours via the ‘invite a friend’ mechanism.
  – After accumulating 20 campus hours, participants may invite up to 10 friends to join the experiment and thereby receive bonus eligible hours.
  – For every friend they invite, after the friend collected 20 campus hours, the inviting participant receives 5 additional eligible hours.
  – The mechanism for inviting a friend is by generating a 6 digit invite code and asking the friend to enter that code.
  – Invited friends will receive 5 eligible hours when they sign up.
- Each phase with the exception of phase 3, has a prize draw at the end of the phase. Additionally, in place of the phase 3 prize draw, which was voided due to lockdown, there is a special prize draw (see below)
- In each prize draw (for phases 1, 2, 4, and 5), prizes are drawn as follows:
  – The chance of winning a prize is based on the eligible hours divided by the total number of eligible hours of all participants.
  – There are 9 prizes in each draw. A single top prize (iPad Pro), 3 second-tier prizes (Android mobile phones), and 5 third-tier prizes (FitBit tracker).
  – A participant may win at most one prize in a draw and this works as follows. First, everyone competes for the top prize and the winner is removed from the pool. Then all remaining participants compete for the second-tier prizes, each time removing the winner. Similarly for the third-tier.
  – A participant is eligible to win at most a single prize from each tier over the course of the experiment. For instance, a winner of the top prize in the first draw will be excluded from winning the top prize in draws that follow (but may still win other prizes).
- Participants can track both their collected campus hours and eligible hours per phase and see how many hours they have collected relative to the distribution of hours collected by other participants (see Fig 3, bottom left plot for a snapshot of the participants leader board showing this distribution).
- All participants are emailed a full report from the prize draw and winners collect directly from experiment staff.
- Winners are asked if they wish to have their picture and short bio posted. This is optional.
- There is a special prize draw in place of phase 3 during the period between phase 3 and phase 4. In this prize draw 10 participants have a chance to win a Fitbit tracker. The prize draw is performed by uniformly selecting 10 winners among all those participants who have accumulated 5 or more campus hours during the period of phase 2.

## Appendix 2: App software

The Safe Blues Android app is based on the Trace Together Android App (Open Trace) [1] and was forked from the Open Trace Android version in April, 2020. The Open Trace software is designed for contact tracing, however we modified it to support the Safe Blues protocols. The app is written in the Kotlin language and has been made available on Google Play since March 2021.

The app is now specifically tailored for the Safe Blues campus experiment and includes on–boarding screens that assign the app instance a unique 10 digit ID. There is no email or other authentication information queried in the app. With the exception of the basic on–boarding screens, there is no user engagement in the app as it runs in the background. The app requires the user to enable location services and Bluetooth. Bluetooth is clearly needed for strand propagation. Location services are not directly part of the Safe Blues system but are needed to recognize that participants are in the geofenced area for the purpose of allocating prizes.

The initial phase of the experiment (see Table 1 in Appendix 6) included the release of several versions of the app that were automatically updated by participants. The sequence of these versions fixed several initial bugs. The most notable bug was a non-random initialization of the random number generator in the app, which caused all participating phones to seed Safe Blues strands in unison. Specifically, in strand batch 1.01 all participating phones decided together whether to seed a strand or not. Once this bug was fixed, a new app version was deployed. This deployment process included the strand batch 1.02 which did not include any strands per-se.

Due to limitations on control over the underlying Bluetooth hardware, the app transfers information between two phones by pairing them together for a brief period of time. We call each such Bluetooth interaction a “ping”. Each time two phones ping each other, they transmit their set of infectious strands as well as the strength of their transmitter. While the app is running, it continuously tries to ping other phones that are part of the experiment, including phones that have been pinged recently. These pings are then combined together into longer Safe Blues sessions (capped at 30 minutes) after which the Safe Blues simulation step runs. The original rationale for the session concept was to allow more flexibility in setting the infection mechanics of the simulation: our system could, for instance, be used to investigate the effect of a perfect contact tracing regime in tandem with these virtual pandemics (where a contact is traced if they spend a minimum duration at a minimum distance with a contact). Furthermore, the underlying Bluetooth systems are far from ideal for this, exhibiting congestion in large groups of participants and being unreliable even when there are few participants. The session system accommodates for some of this unreliability while also giving more accurate estimates of distance.

## Appendix 3: ADS and PMS servers technical details

The Safe Blues experiment system separates the anonymous data server (ADS) and the participant management system (PMS). The ADS is part of the operational Safe Blues system and is used to push new strands to the client apps and record phone strand information through anonymized client IDs generated within the app. The PMS is a system designed specifically for the experiment and is primarily aimed at participant management and prizes. It records how long the app was running in the background while the participant was within the experiment geofence. The PMS accomplishes this by using a separate “experiment ID” that is Completely isolated from the client ID in the ADS. Both systems run in parallel on the phone: the ADS is part of the experiment, while the PMS simply checks whether the participant is on campus with the required permissions and Bluetooth turned on, and relays summary counts to the PMS server.

We chose to physically separate these two servers in order to provide an extra layer of privacy protection for participants as well as to enable future Safe Blues deployments (perhaps operational) to use the ADS while not the PMS. In this separation, the PMS is hosted on a Nectar Research Cloud server managed by The University of Auckland, whilst the ADS is hosted on the commercial Amazon Web Services (AWS) Cloud.

The ADS uses a PostgreSQL database and exposes a gRPC API for phones indicating strand (and virtual social distancing) information, as well as a RESTful admin API. Phones running the app send messages to the ADS every 15 minutes, informing it of the status of the strands. The following are the protocol buffer messages about strands:

~~~
message Strand {
  string name = 13;
  int64 strand_id = 1;
  google.protobuf.Timestamp start_time = 2;
  google.protobuf.Timestamp end_time = 3;
  double seeding_probability = 4;
  // the two parameters of the infection probability map
  double infection_probability_map_p = 5; // strength
  double infection_probability_map_k = 6; // radius
  double infection_probability_map_l = 7; // unused
  // mean and shape of the gamma distribution for incubation period
  double incubation_period_mean_sec = 8;
  double incubation_period_shape = 9;
  // mean and shape of the gamma distribution for infectious period
  double infectious_period_mean_sec = 10;
  double infectious_period_shape = 11;
  uint32 minimum_app_version = 12;
}
~~~

Messages from phones to the ADS are encoded as follows:

~~~
message InfectionReport {
  string client_id = 1;
  int32 version_code = 5;
  repeated int64 current_incubating_strands = 2;
  repeated int64 current_infected_strands = 3;
  repeated int64 current_removed_strands = 4;
}
~~~

All such incoming messages are stored on the ADS Postgres database. Phones are only identified by a temporary 256-bit client ID that changes every 24 hours. Appendix 4 describes the algorithm running on the ADS for interpolation and imputation. Note that the ADS is not aware of the 10 digit participant ID which was only created for the purposes of the experiment.

The PMS stores a list of email addresses associated with each participant ID but does not store any further personal participant information (that is, we do not keep home addresses, names, afflictions, or other private information). The PMS receives messages from the phones via a restful JSON API indicating time spent on campus. An example of such a message is below (where duration and count_active are in units of 15 minute intervals, and the truncated_entry_time is the UNIX timestamp of the entry into campus. As can be seen, these messages indicate the time on campus. The phone generates such a message whenever leaving the geofenced area.

~~~
{
 “participant_id”: 1234567890,
 “version_code”: 60,
 “statuses”: [
   {
     “status_id”: 112,
     “truncated_entry_time”: 18731,
     “duration”: 12,
     “count_active”: 11
  },
   {
     “status_id”: 113,
     “truncated_entry_time”: 18944,
     “duration”: 17,
     “count_active”: 17
  },
   …
]
}
~~~

As shown in the bottom left image of Fig 3, the PMS aggregates these messages in a MySQL database, which is then queried for prize information and for presenting users with their current leader-board standings. The PMS also acts as a web server for a React-based website used for experiment registration, the ‘invite a friend’ mechanism, and the aforementioned leader-board.

Some specific PMS information is made available in accordance with the ethics approval. This is the total number of participants on campus and the total number of registered participants, as shown in Fig 6 (left plot), as well as summary of the distribution of daily campus hours of participants using the mean and a 5-number summary (see the right plot in Fig 6).

## Appendix 4: Interpolation and imputation algorithm

The epidemiological status of strands is stored in the ADS as a collection of reports received from participants’ smartphones. The interpolation and imputation problem is to use these reports to reconstruct each participant’s infection status at arbitrary times throughout the entire day. Specifically, the way in which the experimental data is collected and stored results in two issues that must be resolved: (a) a participant’s status before any reports were received and (b) a participant’s status during the time between reports.

Here, in describing the method used to solve the aforementioned problems, we fix a 24-hour UTC interval, an active strand, and a 256-bit ID. Note that, because a participant’s anonymity 256-bit ID changes every 24 hours, our procedure cannot make use of any information before this window. We collect from the ADS database the following information: the time at which the participant sends their first report, *T*_*F*_ ; the time at which the participant first reported a state of “exposed”, *T*_*E*_; the time at which the participant first reported a state of “infected”, *T*_*I*_ ; and the time at which the participant first reported a state of “recovered”, *T*_*R*_. We choose *T*_*R*_ = ∞ if a “recovered” report is never received, *T*_*I*_ = *T*_*R*_ if an “infected” report is never received, and *T*_*E*_ = *T*_*I*_ if an “exposed” report is never received. Moreover, given any *T* ∈ {*T*_*E*_, *T*_*I*_, *T*_*R*_} with *T* = *T*_*F*_, we set *T* = 0. The participant’s state at an arbitrary time *t* during the day is given by

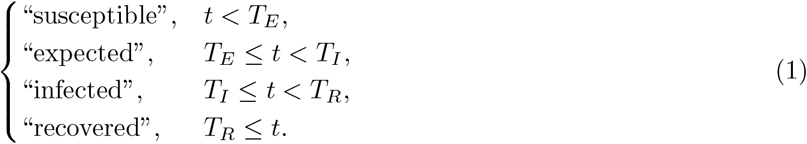

## Appendix 5: Strand details

Here we overview the main purpose of the batches of experiment strands at different stages of the experiment. Specific strands in each batch can be identified from the `strand_id` parameter listed in the `strand.csv` file in the Safe Blues Experiment Data repository (see also Appendix 6). Table 1 details the total number of strands released for each batch during phases 1–3 of the experiment, as well as the strand IDs associated with them.

**Table 1:** Batches of strands and their main purpose in the campus experiment.

| Phase | Batch | No. of strands | Strand IDs | Type | Main Purpose |
| --- | --- | --- | --- | --- | --- |
| 0 | 0.01 | 50 | 1–50 | SEIR, SIR, SEI | The `debug` version, which is the first test of system in a workshop involving 10 users. |
| 1 | 1.01 | 162 | 51–212 | SEIR, SEI, SI | First time testing the system on the experiment population. A bug (“same seed”) was detected. |
| 1 | 1.02 | 1 | 213 | SI | To test the system after fixing the “same seed” bug. |
| 1 | 1.03 | 10 | 214–223 | SEI | After the “same seed” bug, this was an intermediate debug batch while waiting for Google Play app update. |
| 1 | 1.04 | 162 | 224–385 | SEIR, SEI, SI | Same structure of strands as 1.01 released after fixing the “same seed” bug. |
| 1 | 1.05 | 30 | 386–415 | SI | Experimentation of the effect of strength and maximal infection distance on infection via SI strands. |
| 1 | 1.06 | 90 | 416–505 | SI | Further experimentation of the effect of maximal infection distance on infection via SI strands. |
| 1 | 1.07 | 60 | 506–565 | SI | Testing the virtual social distancing feature. |
| 2 | 2.01 | 600 | 566–1165 | SEIR, SIR, SEI, SI | Main release of all types of strands to begin Phase 2 of the experiment. These strands were planned to live until the end of Phase 3. |
| 2 | 2.02 – 2.06 | 600 (per batch) | 1166–4023 | SEIR, SIR, SEI, SI | Weekly releases identical to 2.01 during the first month of the lockdown. |
| 3 | 3.01 – 3.22 | 6 (per batch) | 4024–4155 | SI | As Auckland is still in lockdown, releasing 6 strands per week until the planned start of Phase 4 in case of any changes in behavior over this period |
| 4 & 5 | - | - | - | - | Strands for phases 4 and 5 will be similar to 2.01, currently planned with a release of new strands every two weeks during those phases. |

We now overview the batches of Table 1 starting with the `debug` batch, 0.01, and up to the Phase 3 final batch, `3.22`. Batch `0.01` or the `debug` batch was used as an initial test of the app prior to deploying it to the experimental population. This test trial involved 10 people who attended a Julia language meetup at The University of Queensland on April 21, 2021. This batch contains 50 strands in total, with a combination of SEIR, SIR, and SEI types. Following this, batch `1.01` was the first experimental test of the system as part of The University of Auckland experiment. Immediately after release of this batch, we observed that all participating devices were initialized with the same random number generator seed. This bug, named as the “same seed”, caused all phones to make the same decision about seeding a given strand or not. The bug was fixed and a single SI type strand was released as batch `1.02` to test this. While waiting for a Google Play update for the new app version, strands from batch `1.03` were released as an intermediate step for further testing. These were 10 SEI type strands. Once the bulk of the experiment participants updated their app to the new version, batch `1.04` was released with the same composition of strands initially intended in `1.01`. This batch, `1.04`, constitutes the main experimental batch within Phase 1.

The remaining batches of Phase 1, `1.05 – 1.07`, were aimed at further calibration of the strength and maximal infection distance parameters (as described in Eq (2)), as well as testing the virtual social distancing feature (see Materials and methods section in the main document). Specifically, `1.05` tested a range of distance and strength parameters and their effects on virtual viral spread, and `1.06` refined the search grid in the maximal distance parameters. We were able to see a clear effect of the maximal infection distance parameter on viral spread in both cases (see Fig 9). However, we were unable to detect a significant effect of the strength parameter within this parameter search. We released strands in batch `1.07` at the beginning of the week (Monday morning New Zealand time), and by Wednesday we had inflicted various levels of virtual social distancing, with factors ranging from 1.25 to 3.0. Measurements from this batch clearly indicated that the virtual social distancing mechanism has a significant impact on strand transmission (see Fig 9).

Moving onto the batches of phases 2 and Phase 3, the major batch providing data to date is `2.01`. This batch was released one week into Phase 2 (on Thursday, July 29) and included an extensive variety of strand types based on experience gained in Phase 1. Three weeks after the release, the major New Zealand lockdown took place and this immediately affected strand propagation (see Fig 10 presenting all strands from this batch). With anticipation of the lockdown potentially lifting, we released batches `2.02 – 2.06` weekly where each such batch contains the same types of strands as `2.01`. These batches have not yielded meaningful infections due to the continued lockdown. Finally, throughout Phase 3, and the interim period between Phase 3 and Phase 4, we released the weekly batches `3.01 – 3.22`. These batches, each with only 6 SI strands, are intended to “keep alive” the Safe Blues system.

## Appendix 6: Data structure

The Safe Blues dataset is organized in the `data` repository as shown in Fig 1. The strand parameters determine the epidemiological behavior of each virtual virus–like token, including its transmissibility, incubation duration, and infection duration. A table of these parameters is stored in `data/strands.csv`. This table contains a row for each strand circulated during the campus experiment and contains columns described in Table 1. The transmission data provides aggregate measurements of the spread of strands throughout their reporting periods. We store the progression of the i^th^ strand (`strand_id = i`) as a daily and hourly time series in `strand(i).csv` file. These tables have a row for each time point (either daily or hourly) and have the columns described in Table 2. The participant data presents aggregate information on the engagement of participants throughout the course of the experiment. This is also available as either a daily or hourly time series in `participants.csv` files. Again, these tables contain a row for each time point (either daily or hourly) and contain the columns described in Table 3.

**Figure 1:**
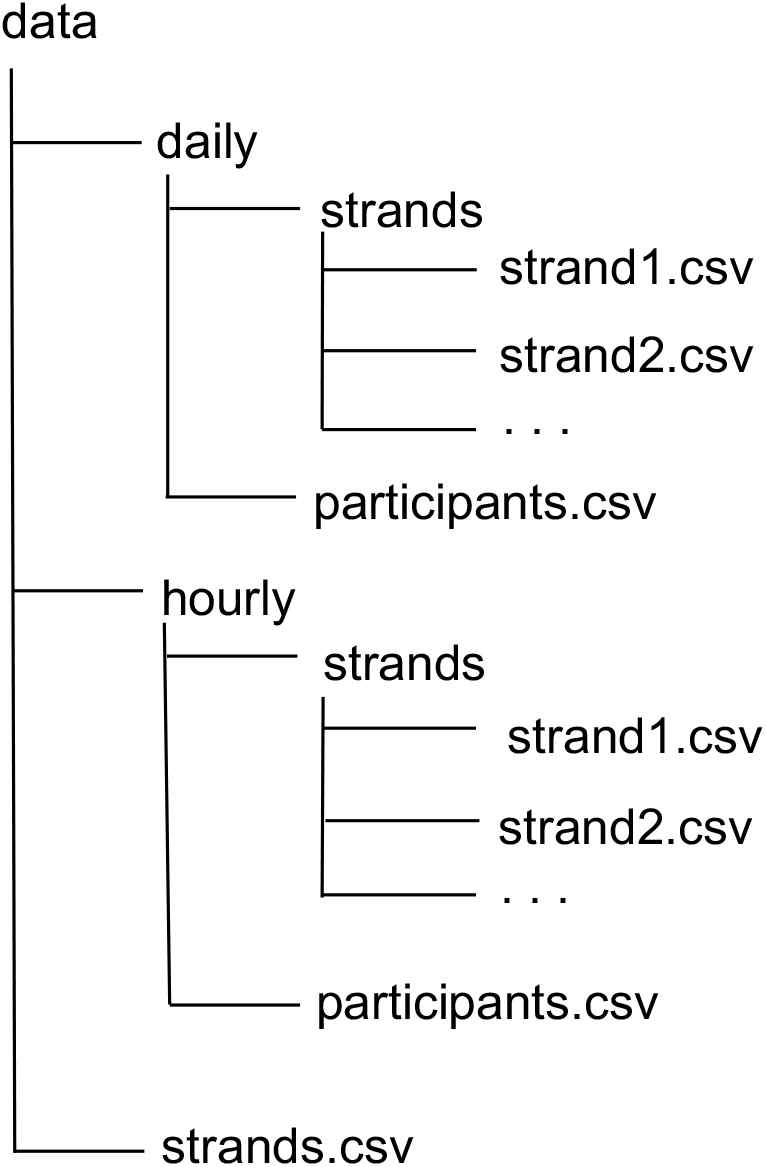
Safe Blues Data Structure.

**Table 1:** Description of the columns in `strands.csv`. The `incubation_mean` and `incubation_shape` values are missing when the `model` is either SI or SIR type. The `infection_mean` and `infection_shape` values are missing when the `model` is either SI or SEI type.

| Name | Description |
| --- | --- |
| **strand_id** | The unique numerical identifier given to each strand |
| **batch** | The release batch or group of strands within which each strand |
| **model** | The epidemiological model used by each strand (either SI, SEI, SIR, or SEIR) |
| **start_utc** | The time at which each strand's reporting begins in UTC timezone |
| **start_nzt** | The time at which each strand's reporting begins in NZST/NZDT timezone |
| **seed_utc** | The time at which each strand's initial infections occur in UTC timezone |
| **seed_nzt** | The time at which each strand's initial infections occur in NZST/NZDT timezone |
| **stop_utc** | The time at which each strand's reporting ends in UTC timezone |
| **stop_nzt** | The time at which each strand's reporting ends in NZST/NZDT timezone |
| **initial** | A participant's probability of initial infection for each strand |
| **strength** | The strength of transmission for each strand |
| **radius** | The maximal infection distance of transmission for each strand |
| **incubation_mean** | The mean parameter for each strand's gamma distributed incubation duration |
| **incubation_shape** | The shape parameter for each strand's gamma distributed incubation duration |
| **infection_mean** | The mean parameters of each strand's gamma distributed infection duration |
| **infection_shape** | The shape parameters of each strand's gamma distributed infection duration |

**Table 2:** Description of the columns in the `strand_i.csv` file, where `i` is the `i`^th^ strand. The values for `exposed` column are missing when the `model` is either SI or SIR type. The values for `recovered` column are missing when the `model` is either SI or SEI type.

| Name | Description |
| --- | --- |
| **strand_id** | The unique numerical identifier given to each strand |
| **time_utc** | The time of each measurement in UTC timezone |
| **time_nzt** | The time of each measurement in NZST/NZDT timezone |
| **susceptible** | The current number of participants who are susceptible to the strand |
| **exposed** | The current number of participants who are incubating the strand |
| **infected** | The current number of participants who are infected with the strand |
| **recovered** | The current number of participants who have recovered from the strand |
| **distance_factor** | The artificial distance multiplier used to emulate social distancing |

**Table 3:** Description of the columns in the `participants.csv` files. All statistics reported in the two `participants.csv` files are in reference to NZST/NZDT days except for `count_reporting`, which is in reference to UTC days. The mean and 5 number summary correspond to the campus hours collected by participants who attended campus on the current day. Values for the mean and 5 number summary are missing when `count_campus` is less than or equal to 5.

| Name | Description |
| --- | --- |
| `time_utc` | The time of each measurement in UTC timezone |
| `time_nzt` | The time of each measurement in NZST/NZDT timezone |
| `count_campus` | The number of participants on campus during the current day |
| `count_reporting` | The number of participants whose phones are sending reports during the current UTC day |
| `count_registered` | The number of participants who have registered for the experiment by the current day |
| `hours_mean` | The mean number of campus hours |
| `hours_min` | The minimum number of campus hours collected |
| `hours_q1` | The lower quartile number of campus hours collected |
| `hours_q2` | The median number of campus hours collected |
| `hours_q3` | The upper quartile number of campus hours collected |
| `hours_max` | The maximum number of campus hours collected |

